# Visual Cortical Thickness Increases with Prolonged Artificial Vision Restoration

**DOI:** 10.1101/2024.06.26.24309493

**Authors:** Noelle R. B. Stiles, Jeiran Choupan, Hossein Ameri, Vivek R. Patel, Yonggang Shi

## Abstract

The Argus II retinal prosthesis restores visual perception to late blind patients. It has been shown that structural changes occur in the brain due to late-onset blindness, including cortical thinning in visual regions of the brain. Following vision restoration, it is not yet known whether these visual regions are reinvigorated and regain a normal cortical thickness or retain the diminished thickness from blindness.

We evaluated the cortical thicknesses of ten Argus II Retinal Prostheses patients, ten blind patients, and thirteen sighted participants. The Argus II patients on average had a thicker left Cuneus Cortex and Lateral Occipital Cortex relative to the blind patients. The duration of the Argus II use (time since implant in active users) significantly partially correlated with thicker visual cortical regions in the left hemisphere. Furthermore, in the two case studies (scanned before and after implantation), the patient with longer device use (44.5 months) had an increase in the cortical thickness of visual regions, whereas the shorter-using patient did not (6.5 months). Finally, a third case, scanned at three time points post-implantation, showed an increase in cortical thickness in the Lateral Occipital Cortex between 43.5 and 57 months, which was maintained even after 3 years of disuse (106 months).

Overall, the Argus II patients’ cortical thickness was on average significantly rejuvenated in two higher visual regions and, patients using the implant for a longer duration had thicker visual regions. This research raises the possibility of structural plasticity reversing visual cortical atrophy in late-blind patients with prolonged vision restoration.

## Introduction

For most of the 20^th^ century extensive and complex damage to retinal cells, vasculature, and neural networks prevented the restoration of visual perception to those blinded by degenerative retinal disease. However, modern advances in biomedical engineering as well as molecular, and cell biology developed over the last few decades have facilitated the innovation of emerging therapeutics and implants to restore functional vision to the visually impaired. These therapeutics and implants include biomedical devices such as retinal and cortical prostheses (Beauchamp and others 2020; Humayun and others 2012; Luo and others 2016; Luo and Da Cruz 2014; Luo and da Cruz 2016; Niketeghad and Pouratian 2019; Weiland and others 2011; Zhou and others 2013; Zrenner 2013; Zrenner and others 2011), as well as biomolecular therapies, such as gene therapies (Apte 2018; Lam and others 2019; Mowad and others 2020), optogenetic therapies (Baker and Flannery 2018; Ferrari and others 2020; Simunovic and others 2019), and stem cell therapies (Kashani and others 2018; Roska and Sahel 2018). Retinal prostheses have been one of the approaches most studied and realized, with hundreds of patients already implanted with prostheses worldwide.

Retinal prostheses restore low-resolution visual perception to the blind by stimulating viable retinal cells with a video stream from a head-mounted camera (Figure 1) (Luo and da Cruz 2016; Zhou and others 2013). In this paper we study the Argus II retinal prosthesis, which was manufactured by Second Sight Medical Products and has been approved by the FDA for implantation in one eye of Retinitis Pigmentosa (RP) patients with light perception or less. The Argus II device has been implanted in approximately 350 visually impaired patients worldwide (2019).

**Figure 1:**
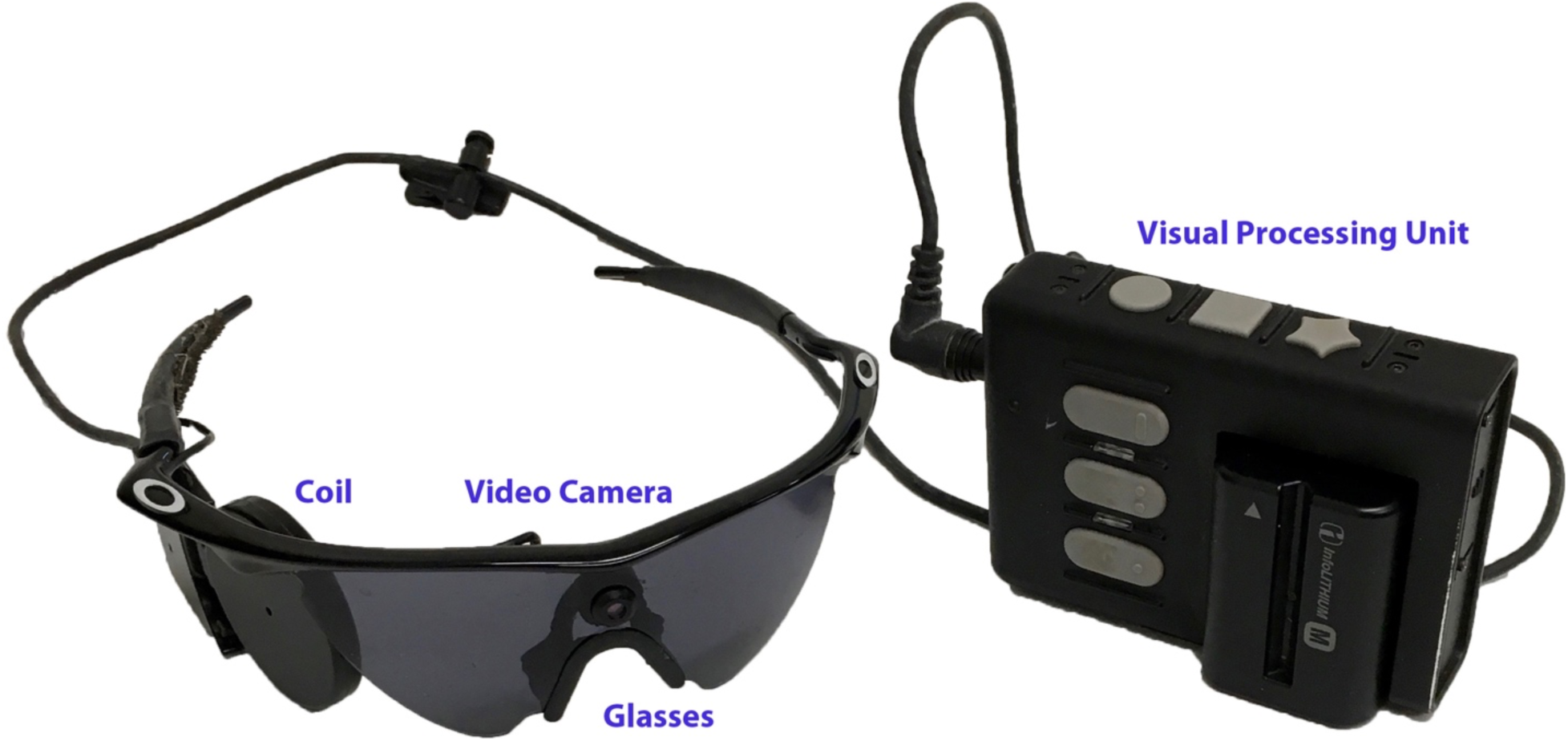
Argus II Device External Components. An image of the Argus II retinal prosthesis external components including the pair of glasses with a head mounted camera, and the visual processing unit. The implanted internal components are not included in this image.

The Argus II device consists of 60 electrodes (60 pixels), which generate ultra-low resolution vision with the ability to determine the direction of motion, match auditory and visual stimuli, identify basic shapes, and perform object localization (Humayun and others 2012; Kotecha and others 2014; Luo and others 2014; Stiles and others 2021; Stronks and Dagnelie 2014). However, Argus II artificial visual perception is quite different than natural vison. In particular, prosthesis-driven perception is highly variable across the patient population and can require substantial effort and attention to perform basic tasks at a slow rate (relative to low vision and sighted individuals) (Luo and others 2016; Luo and da Cruz 2016; Zhou and others 2013). These challenges could be due to the elongated and curved shape of the phosphenes (pixels) generated by electrical stimulation (Beyeler and others 2019), perceptual fading (Avraham and others 2021; Fornos and others 2012), deterioration of retinal circuitry during blindness (Marc and Jones 2003), the abnormal stimulation of retinal ganglion cells by electrodes (Beyeler and others 2019), and/or visual cortical changes during blindness (Pascual-Leone and Hamilton 2001; Sadato and others 2004). The cortical changes during blindness can include both functional changes, such as the crossmodal repurposing of dormant visual regions with auditory or tactile processing (Amedi and others 2005; Amedi and others 2007; Sadato and others 2004), and structural changes, such as the atrophy of visual white matter pathways and grey matter cortical thicknesses (Burge and others 2016; Leporé and others 2010; Machado and others 2017). Both the functional and structural changes of the visual cortical neural network during decades of late blindness may limit the behavioral outcomes of Argus II patients. In addition, the ability for the visual cortex to reorganize and adapt to restored visual input may have implications for Argus II patient outcomes.

In this paper, we investigate the structural changes of the grey matter cortical thicknesses in Argus II retinal prosthesis patients with late-stage Retinitis Pigmentosa (RP). Late-stage RP causes at a minimum severe constriction of peripheral vision; however, it can also progress to complete loss of visual perception with only light perception remaining (as with the Argus II device candidates). Late-stage RP patients, similar to most late blind patients, have been shown to have significant changes to the structure of the visual cortex due to vision loss (Castaldi and others 2019; Cunningham and others 2015a; Ferreira and others 2017; Machado and others 2017; Sanda and others 2018). In particular, late-stage RP patients have been found to have significant thinning of primary visual cortex when compared to sighted controls (Sanda and others 2018), which is hypothesized to be due to atrophy that progresses from the damaged retina, extending through the visual pathway to visual cortical regions (i.e. trans-synaptic neurodegeneration, or Wallerian degeneration) (Machado and others 2017). RP patients have also been shown to have a reduction in visual cortex total grey matter that negatively correlates with visual function. In other words, patients with more severe vision loss were found to have more grey matter loss in visual cortical regions (Machado and others 2017).

In this study we investigate whether vision restoration by prostheses such as the Argus II device can reverse the cortical structural atrophy generated by an extended period of blindness due to late-stage RP. We evaluate this question by comparing the visual cortical thickness of ten Argus II retinal prosthesis patients to ten late-stage RP patients (blind controls) and thirteen sighted controls (both age-matched). We will also evaluate the visual cortical thicknesses before and after implantation in two Argus II case studies, and at multiple timepoints following implantation in a third Argus II case study.

## Materials and Methods

### Participants

Ten Argus II patients (7 male, 3 female) with an average age of 61 years old participated in this research study at the University of Southern California (USC) (Table 1). The Argus II patients had the device implanted for an average of 51.60 months (4.30 years), and experienced substantial vision loss for an average of 32.22 years (Note: One patient did not report their duration of blindness). All of the Argus II patients had light perception or less in both eyes when the Argus II device was turned off, and the Argus II device was implanted in the left eye in 3 patients, and the right eye in 7 patients.

**Table 1.**
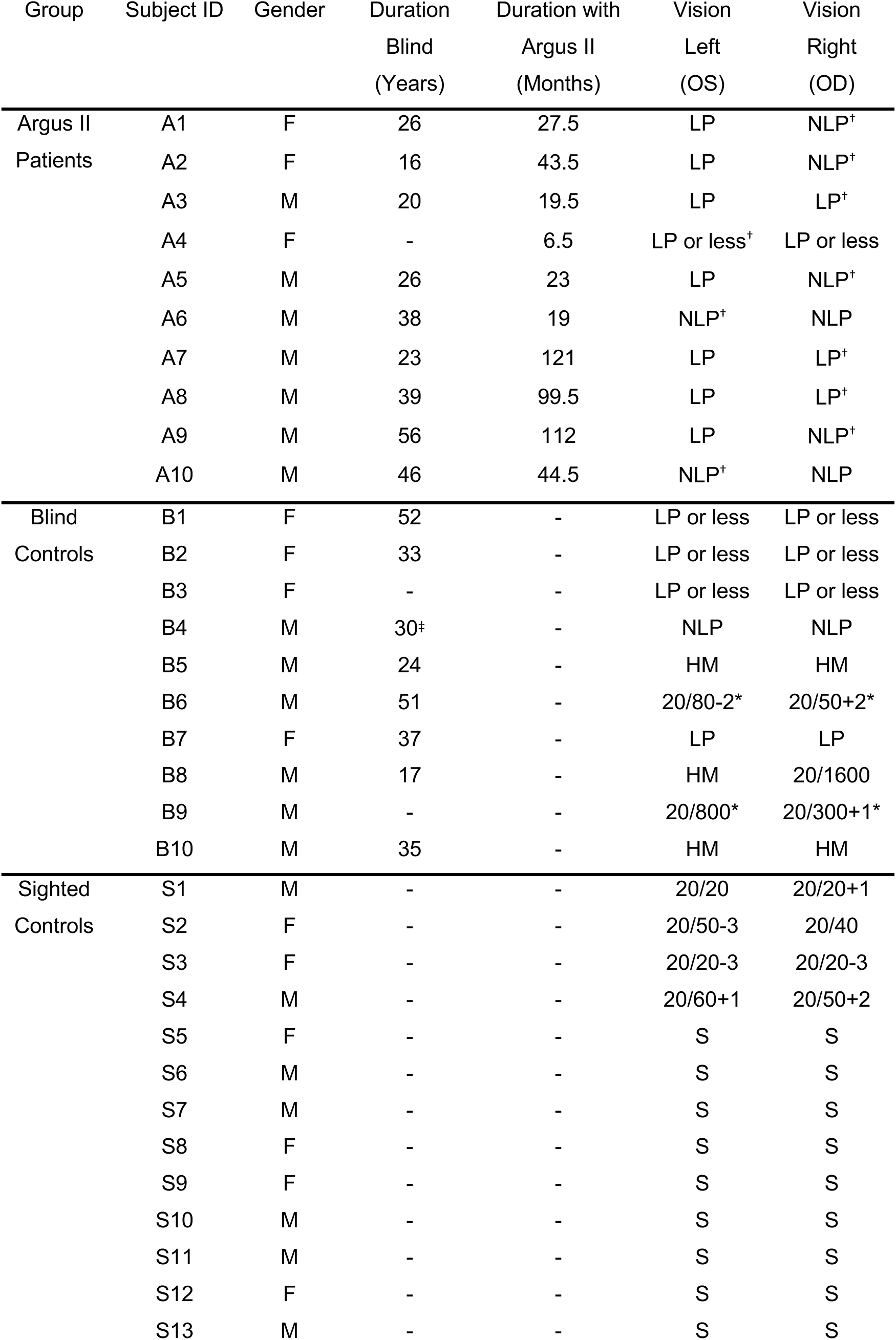
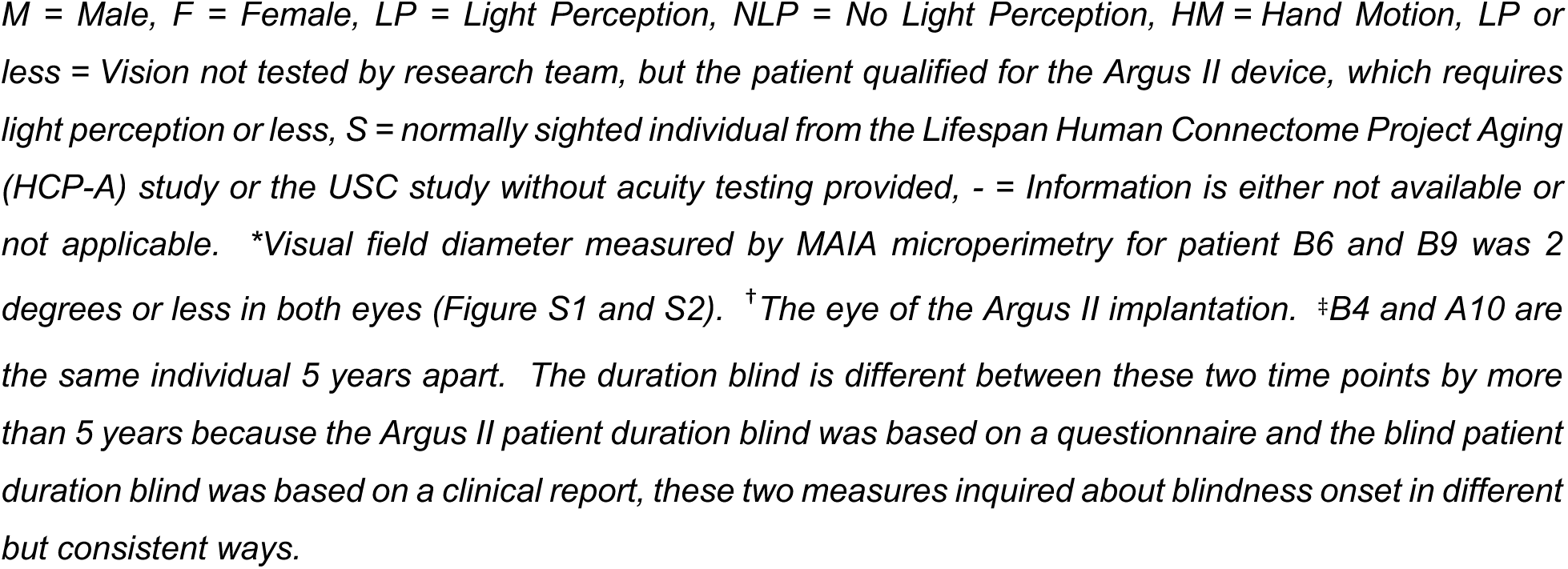
Argus II Patients, Blind Controls, and Sighted Controls Demographic Information.

| Group | Subject ID | Gender | Duration<br>Blind<br>(Years) | Duration with<br>Argus II<br>(Months) | Vision<br>Left<br>(OS) | Vision<br>Right<br>(OD) |
| --- | --- | --- | --- | --- | --- | --- |
| Argus II<br>Patients | A1 | F | 26 | 27.5 | LP | NLP<sup>†</sup> |
|  | A2 | F | 16 | 43.5 | LP | NLP<sup>†</sup> |
|  | A3 | M | 20 | 19.5 | LP | LP<sup>†</sup> |
|  | A4 | F | - | 6.5 | LP or less<sup>†</sup> | LP or less |
|  | A5 | M | 26 | 23 | LP | NLP<sup>†</sup> |
|  | A6 | M | 38 | 19 | NLP<sup>†</sup> | NLP |
|  | A7 | M | 23 | 121 | LP | LP<sup>†</sup> |
|  | A8 | M | 39 | 99.5 | LP | LP<sup>†</sup> |
|  | A9 | M | 56 | 112 | LP | NLP<sup>†</sup> |
|  | A10 | M | 46 | 44.5 | NLP<sup>†</sup> | NLP |
| Blind<br>Controls | B1 | F | 52 | - | LP or less | LP or less |
|  | B2 | F | 33 | - | LP or less | LP or less |
|  | B3 | F | - | - | LP or less | LP or less |
|  | B4 | M | 30<sup>‡</sup> | - | NLP | NLP |
|  | B5 | M | 24 | - | HM | HM |
|  | B6 | M | 51 | - | 20/80-2* | 20/50+2* |
|  | B7 | F | 37 | - | LP | LP |
|  | B8 | M | 17 | - | HM | 20/1600 |
|  | B9 | M | - | - | 20/800* | 20/300+1* |
|  | B10 | M | 35 | - | HM | HM |
| Sighted<br>Controls | S1 | M | - | - | 20/20 | 20/20+1 |
|  | S2 | F | - | - | 20/50-3 | 20/40 |
|  | S3 | F | - | - | 20/20-3 | 20/20-3 |
|  | S4 | M | - | - | 20/60+1 | 20/50+2 |
|  | S5 | F | - | - | S | S |
|  | S6 | M | - | - | S | S |
|  | S7 | M | - | - | S | S |
|  | S8 | F | - | - | S | S |
|  | S9 | F | - | - | S | S |
|  | S10 | M | - | - | S | S |
|  | S11 | M | - | - | S | S |
|  | S12 | F | - | - | S | S |
|  | S13 | M | - | - | S | S |

Argus II patient A4 and A10 (included in the Argus II patient group) were also scanned before device implantation. This pre-implantation data from patient A4 and A10 are included in the blind patient cohort (detailed below), as patient B3 and B4, respectively. In addition, we also compared the pre-implantation and post-implantation cortical thicknesses for these two patients as two separate case studies in the results section without statistical analyses.

One other Argus II patient also participated in a longitudinal case study post-implantation. Patient A2 was scanned at 43.5, 57, and 106 months after Argus II implantation. At the first two scans she was actively using the Argus II device, however, at the last scan at 106 months, she had not been using the Argus II device for 36 months. The first scan of patient A2 (at 43.5 months) is included in the group level analysis, and a case study comparison between the three scans is also included in the results section without statistical analyses.

Ten advanced RP control patients participated in this research study at USC (Table 1). The advanced RP patients are called the “blind control group” throughout the paper. The blind control group had an average age of 51 years at the time of the experiment and consisted of 6 male and 4 female individuals. The blind control group all had severe vision loss with a range of acuity between no light perception and 20/50+2 (Table 1). The only patient with acuity above 20/300+1 was patient B6, who had a visual acuity of 20/80-2 and 20/50+2, left and right eye respectively. Patient B6 was classified as severe RP and included in the blind patient group because he had a visual field of 2 degrees and a low level of light detection in both eyes as measured with MAIA microperimetry (Figure S1). Patient B9 (visual acuity: 20/800 in the left eye and 20/300+1 in the right eye (Table 1)), also had a visual field of 2 degrees as measured with MAIA microperimetry (Figure S2).

Note: Four of the blind control patients (B1, B2, B3, and B4) qualified for the Argus II implant. Two of these patients received the implant and were scanned after implantation as patient A4 and A10 (detailed above). The other two patients either did not receive the implant or did not return for a second scan following implantation.

Thirteen sighted control individuals participated in this research study. Seven of the sighted controls (participants S1 through S7) had data collected at USC with the same protocol as the Argus II patients and blind control patients detailed above. Due to the small size of the USC sighted cohort, six of the sighted controls (participants S8 through S13) were included from the Lifespan Human Connectome Project Aging (HCP-A) study (Bookheimer and others 2019). The HCP-A participants were chosen from that database to match the age and gender of the first six Argus II patients (in order from S8 through S13 the HCP-A patients included are: HCA61, HCA71, HCA73, HCA76, HCA79, and HCA88). A comparison between the two patient groups (USC sighted controls, and HCP-A sighted controls) did not show a significant difference (Supplemental Information, Figure S3) except for left pericalcarine cortex (*p* = 0.04) and left lingual gyrus (*p* = 0.04). The sighted control group (including participants from USC and HCP-A) had an average age of 55.23 years old and consisted of 7 males and 6 females (Table 1).

The neuroimaging data for the Argus II patients (patients A1 through A10), the blind patients (patients B1 through B10), and seven of the sighted participants (participants S1 through S7) were collected at USC and approved by the USC Institutional Review Board. The participants all gave written informed consent before performing the experiments.

### Argus II Retinal Prosthesis System

The Argus II retinal prosthesis system was manufactured by Second Sight Medical Products. The device consists of an implanted microelectrode array and coil, and the external glasses and visual processing unit (Figure 1). The microelectrode array has 60 electrodes, which electrically stimulate the retinal surface. Visual information is initially captured by a camera on the pair of glasses that sends the video stream to the visual processing unit to convert the video into stimulation parameters. The information is then sent back to the glasses via a cable and communicated into the eye with a glasses-mounted coil, that transmits a signal to an internal coil and then the microelectrode array. The Argus II was approved for patients with Retinitis Pigmentosa and a visual acuity of light perception or less. Outside this indication, it has been implanted in patients with dry age-related macular degeneration in a clinical trial.

Following device implantation and activation, patients are offered rehabilitation training (often in their home environment) to facilitate the learning of basic visual tasks and functions. Seven of the Argus II patients in this study participated in the Second Sight provided training (one patient did not provide this information). Additional information about the training of the Argus II patients is detailed in Table S1. Following directed rehabilitation training, patients can use the device in the tasks of daily living. The Argus II patients scanned were active users of the device, nine patients self-reported their frequency of use, which ranged from every day to once per month (frequency averaged over total time since implant) (Table S1).

### USC and HCP-A MRI Scanning Protocols

The USC MRI scans were performed on a 3-Tesla Siemens Prisma MRI scanner with a 32-channel Siemens head coil. For each participant a T1-weighted anatomical image with magnetization-prepared rapid gradient-echo (MPRAGE) was acquired in axial orientation (TR = 2400 ms, TE = 2.22 ms, flip angle = 8, FOV = 256 × 256 mm^2^, voxel size 0.8 mm isotropic voxels, number of slices = 208, acceleration factor (GRAPPA) = 2). The data collection followed the Human Connectome Project LifeSpan protocol (VD13D). The sighted control data for participants S8 through S13 was derived from the HCP-A database (Bookheimer and others 2019). The HCP-A data was acquired on a 3-Tesla Siemens Prisma MRI scanner with a 32-channel head coil. The T1-weighted anatomical image was collected with multiecho MPRAGE (TR/TI = 2500/1000, TE = 1.8/3.6/5.4/7.2 ms, flip angle = 8, sagittal FOV = 256 × 240 × 166 mm, voxel size 0.8 mm isotropic voxels, 4 echoes per line of k-space, matrix = 320 x 300 x 208 slices, 7.7% slice oversampling, pixel bandwidth = 744 Hz/pixel, acceleration factor (GRAPPA) = 2). Motion-induced re-acquisition were allowed for up to 30 TRs. (Harms and others 2018).

#### MRI Data Processing Details

The T1-weighted structural images were processed in Freesurfer 7.1.1 software using the recon-all function, with a fully automated directive (-all). (Desikan and others 2006; Klein and Tourville 2012). Four visual regions of interest were compared between participant groups (Argus II patients, blind patients, and sighted controls): Pericalcarine Cortex, Cuneus Cortex, Lingual Gyrus, and Lateral Occipital Cortex. The regions of interest were selected to include all occipital lobe regions as delineated by the Desikan-Killiany atlas. The left and right cortical regions were evaluated separately.

The segmentation of the thalamic nuclei was performed with the segmentThalamicNuclei.sh function in Freesurfer 7.1.1. The left and right Lateral Geniculate Nucleus (LGN) volumes were retrieved from the thalamic-nuclei.lh.v12.T1 and the thalamic-nuclei.rh.v12.T1 files respectively. The left and right LGN volumes were normalized by the brain segmentation volume without ventricles (aseg file).

Note: Normalization by each participant’s average cortical thickness or hemispheric brain volume was performed in order to remove the cross-subject variation due to differences in total brain volume. This normalization follows the standard practice in the cortical thickness literature (Aguirre and others 2016; Cunningham and others 2015a; Ferreira and others 2017; Jiang and others 2015; Sanda and others 2018).

### Statistical Analyses and Data Visualization

The cortical thickness data was evaluated for normality using the Shapiro-Wilk and Shapiro-Francia tests (BenSaïda 2014), and failed to be classified as normal. Therefore, the Kruskal-Wallis non-parametric one-way ANOVA analysis was performed for each region of interest. If the one-way ANOVA showed a significant variation between participant groups (*p* < 0.05), group comparisons were made. The individual group pairs were compared using the multcompare function (Fisher’s Least Significant Difference Procedure). The multcompare function based its analysis on a one-way ANOVA for the cortical region of interest comparing the three participant groups (this prevented comparisons between the cortical regions, which was not of interest). The visualization of the Desikan-Killiany atlas was performed in FreeView.

Partial correlations were performed in MATLAB using the partialcorr function. Cortical thickness was correlated with the patient’s duration of prosthesis use (calculated as the period between patient’s self-reported date of implantation and the date of testing), the patient’s duration of blindness (self-reported), or the Argus II patient functionality (shape matching task, methods for this task are detailed in Stiles et al (Stiles and others 2022)) while controlling for patient age.

### Data Availability

Neuroimaging data will be made available through the Connectome Coordination Facility (CCF) (https://www.humanconnectome.org/) as a part of the Human Connectome for Low Vision Project funded by the National Institutes of Health.

## Results

### Group Level Analyses

#### Left Hemisphere

The left Pericalcarine Cortex did not have significant differences among the participant groups (*H* (2, *n* = 33) = 2.13, *p* = 0.34) (Figure 2).

**Figure 2:**
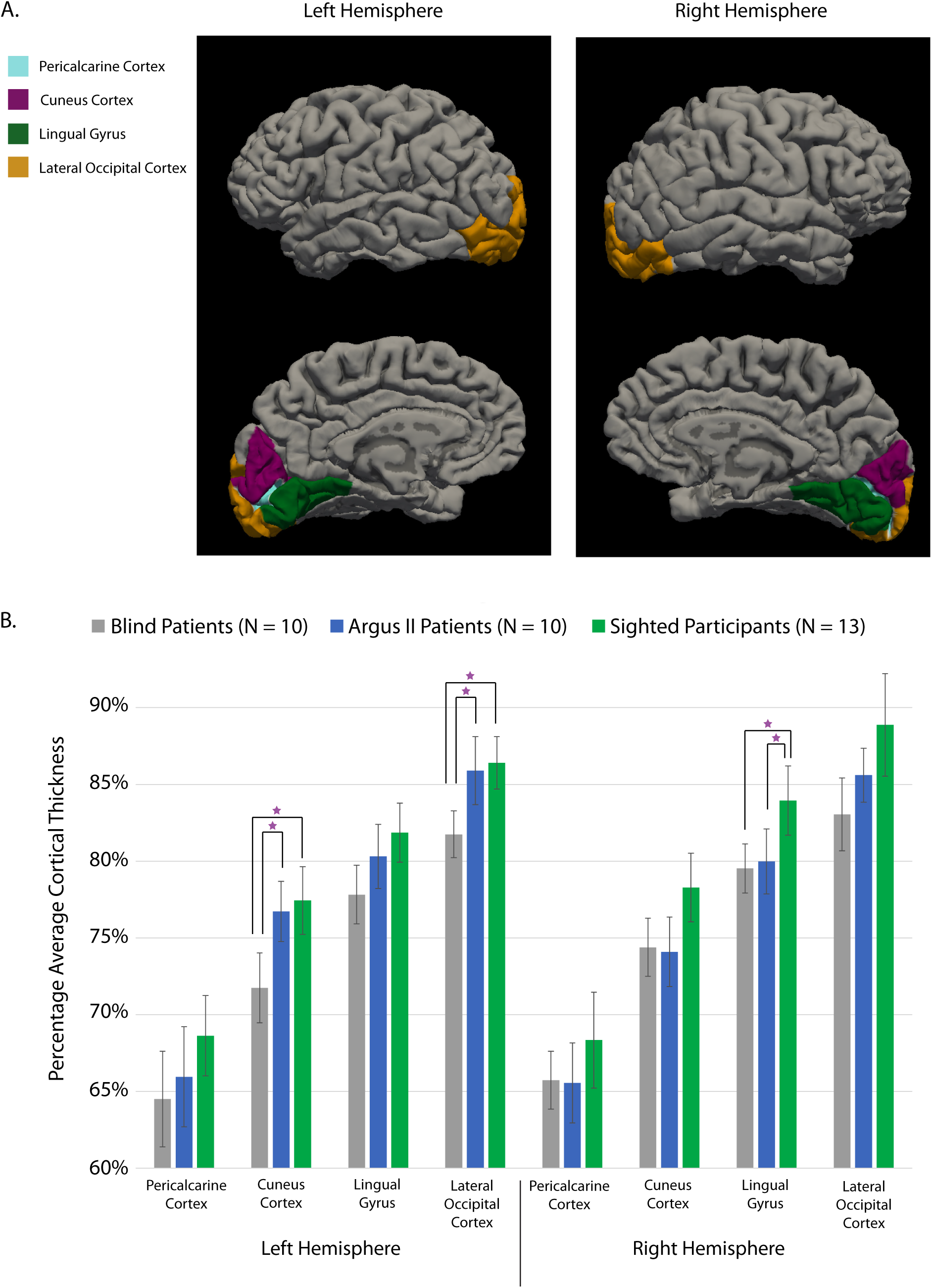
Visual Region Cortical Thicknesses. Panel A shows the parcellation of the occipital regions with the Desikan-Killiany atlas in an example Argus II patient, A1. Note: The Pericalcarine Cortex (light blue) can be found between the Cuneus Cortex and Lingual Gyrus as a sulcus (inward cortical fold). Panel B shows the cortical thicknesses for the four visual areas highlighted in Panel A in the three participant groups: the blind patients, the Argus II patients, and the sighted controls (detailed in Table 1). The cortical thickness measures are normalized by the average cortical thickness in the same hemisphere for each participant. Purple stars indicate significant differences between participant groups (*p* < 0.05). The full length of each error bar represents one standard deviation.

The left Cuneus Cortex had significant variation across the participant groups (*H* (2, *n* = 33) = 7.41, *p* = 0.02). The Argus II patients and the sighted participants had on average a significantly thicker left Cuneus Cortex than the blind patients (Argus II patients vs. Blind controls: *p* = 0.03; Sighted controls vs. Blind controls: *p* = 0.01) (Figure 2). The Argus II patients did not have a significantly different cortical thickness than the sighted controls (*p* = 0.74) (Figure 2).

The left Lingual Gyrus did not have significant differences among the participant groups (*H* (2, *n* = 33) = 5.62, *p* = 0.06) (Figure 2).

The left Lateral Occipital Cortex had significant variation across the participant groups (*H* (2, *n* = 33) = 7.16, *p* = 0.03). The Argus II patients and the sighted participants were found to have a significantly thicker left Lateral Occipital Cortex than the blind patients (Argus II patients vs. Blind controls: *p* = 0.04; Sighted controls vs. Blind controls: *p* = 0.01) (Figure 2). The Argus II patients’ cortical thickness of left Lateral Occipital Cortex was not significantly different than the sighted controls (Argus II patients vs. Sighted controls: *p* = 0.68) (Figure 2).

The left Lateral Geniculate Nucleus volume results are reported in the Supplemental Information, Figure S4.

#### Right Hemisphere

The right hemisphere did not have any significant differences between participant groups in the Pericalcarine Cortex (*H* (2, *n* = 33) = 2.07, *p* = 0.36), Cuneus Cortex (*H* (2, *n* = 33) = 5.22, *p* = 0.07) or Lateral Occipital Cortex (*H* (2, *n* = 33) = 5.09, *p* = 0.08) (Figure 2).

However, the right Lingual Gyrus had significant variation across the participant groups (*H* (2, *n* = 33) = 7.34, *p* = 0.03). The sighted participants had on average a significantly thicker left Cuneus Cortex than the Argus II patient group and the blind control group (Sighted controls vs. Argus II patients: *p* = 0.04; Sighted controls vs. Blind controls: *p* = 0.01) (Figure 2). The Argus II patients’ cortical thickness of right Lingual Gyrus was not significantly different than the blind controls (*p* = 0.75) (Figure 2).

The right Lateral Geniculate Nucleus volume results are reported in the Supplemental Information, Figure S4.

#### Correlations Between Cortical Thickness and Demographics

The duration of prosthesis use (*N* = 10) was significantly positively partially correlated with the average cortical thickness of the four visual regions of interest of the left hemisphere (*rho* = 0.83, *p* = 6.2 x 10^−3^) but not the right hemisphere (*rho* = 0.29, *p* = 0.45)(controlling for age)(Figure 3). To more precisely evaluate this correlation, partial correlation analyses for each visual region in the left hemisphere were also performed. The cortical thickness of the left Pericalcarine Cortex (*rho* = 0.68, *p* = 0.04) and Cuneus Cortex (*rho* = 0.82, *p* = 6.8 x 10^−3^) significantly partially correlated with the duration of prosthesis use (controlling for age).

**Figure 3:**
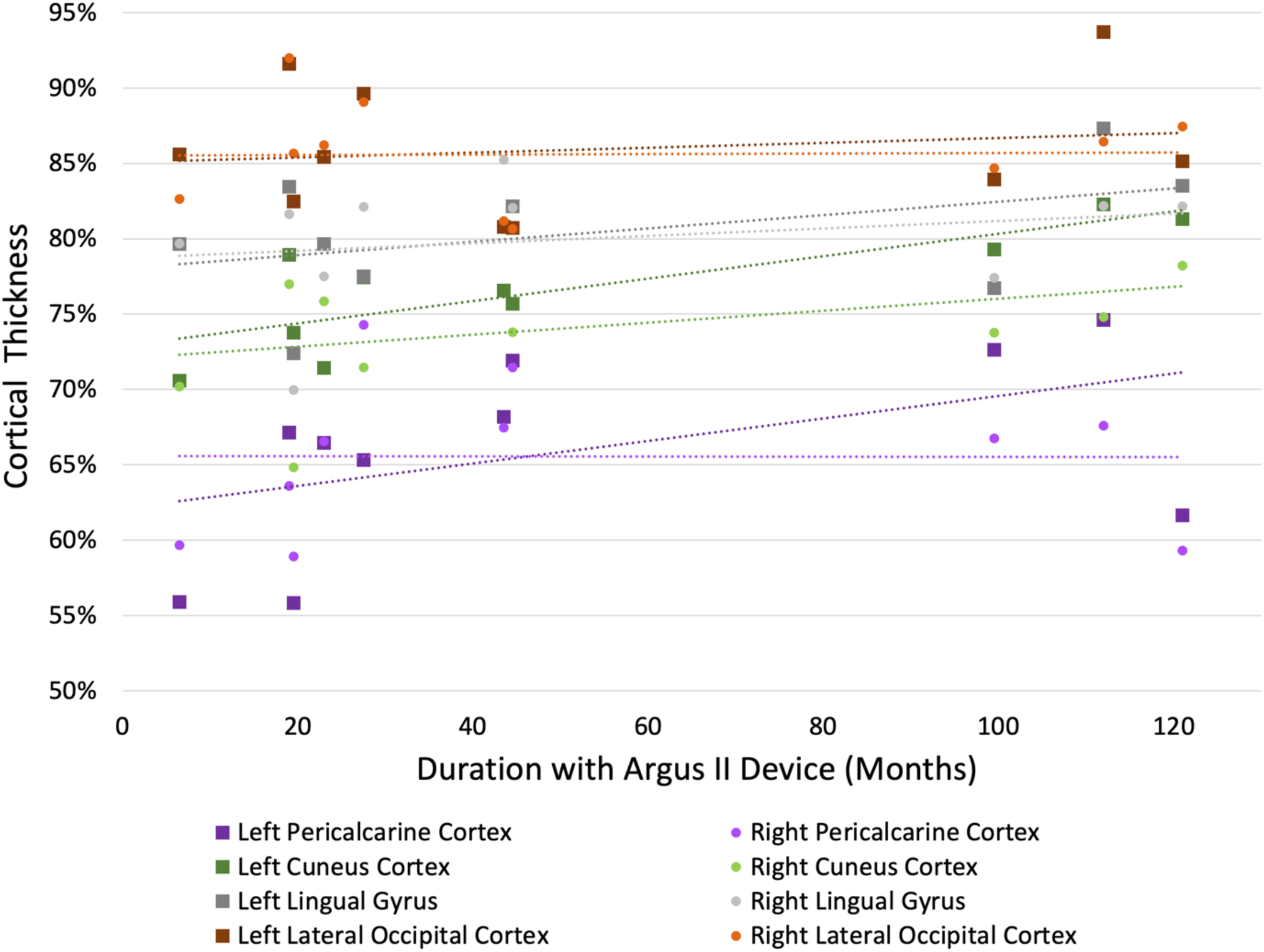
Correlation between Visual Cortical Thickness and the Duration of Argus II Device Use. Figure 3 shows a correlation plot for the Argus II patients (*N* = 10) between the visual cortical thickness and the duration of Argus II device use. Each color represents a different visual region of interest including the left and right hemispheres of the Pericalcarine Cortex, Cuneus Cortex, Lingual Gyrus, and Lateral Occipital Cortex. Each square or dot is a data point for one subject in that visual region of interest.

**Figure 4:**
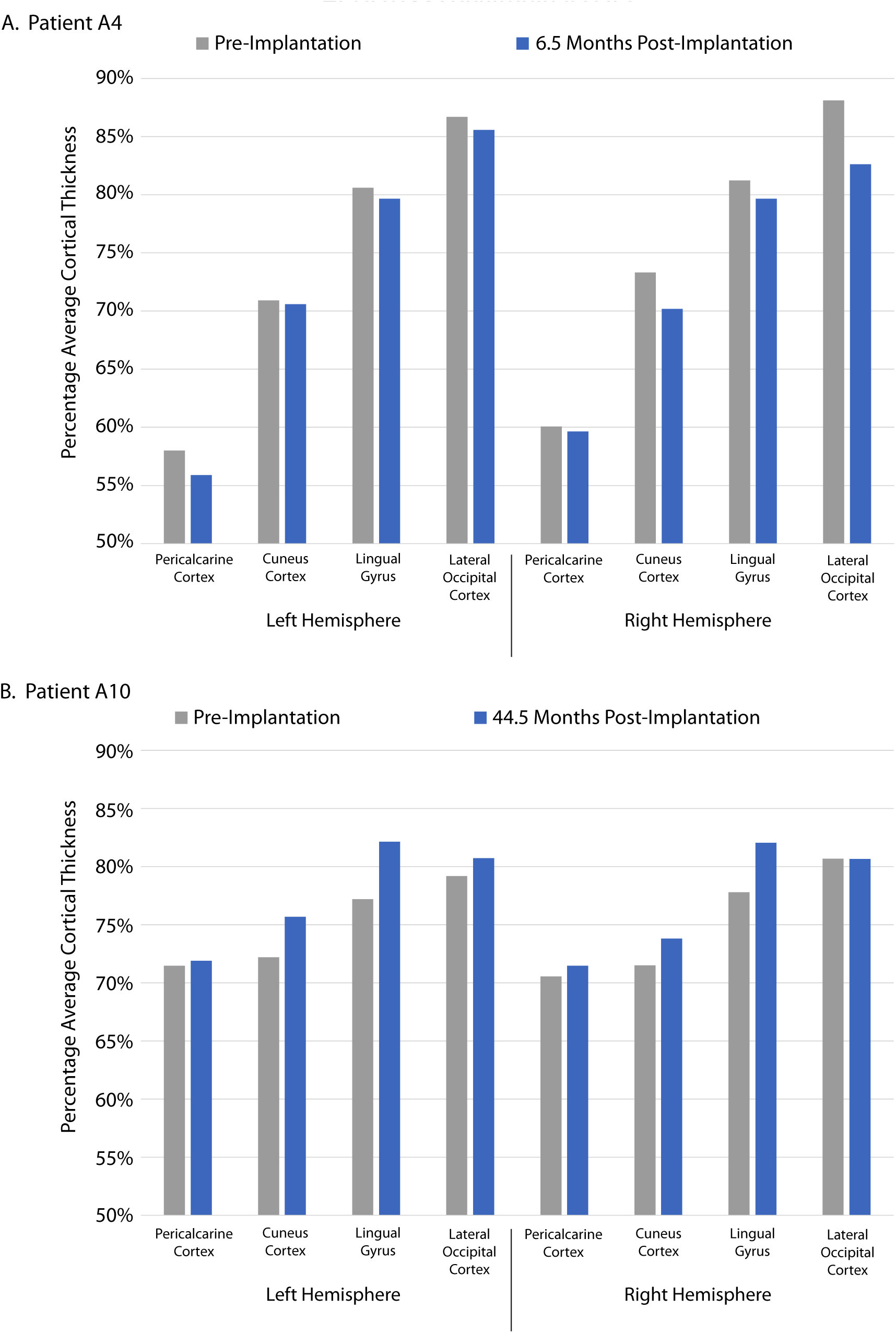
Individual Pre and Post Implantation Visual Cortical Thicknesses. Figure 4 shows the visual cortical thicknesses for two case studies, A4 and A10, which have scans before and after the implantation of the Argus II device (details in Table 1). Patient A4 was scanned 6.5 (0.54 years) after Argus II implantation and Patient A10 was scanned 44.5 months (3.71 years) after Argus II implantation

However, an Argus II functionality measure (shape matching task derived from Stiles et al, detailed in Methods section) and the patient duration of blindness did not partially correlate (controlling for age) with the average cortical thickness of the four visual regions of interest (Shape Task, *N* = 8: left hemisphere: *rho* = –0.13, *p* = 0.77; right hemisphere: *rho* = 0.18, *p* = 0.69)(Duration of blindness, *N* = 9: left hemisphere: *rho* = 0.62, *p* = 0.18; right hemisphere: *rho* = –0.19, *p* = 0.65).

### Individual Longitudinal Analyses

#### Pre and Post Implantation Scans (Two Case Studies)

Patients A4 and A10 were scanned before and after implantation with the Argus II device, permitting a direct within-patient comparison of visual cortical thickness (the left and right LGN volumes are reported in the Supplemental Information, Figure S5). Patient A4 was scanned 6.5 months (0.54 years) following Argus II implantation, and had an average reduction in visual cortical thickness of 1.9 percent relative to the pre-implantation scan (Figure S5A). Patient A10 was scanned 44.5 months (3.71 years) following the Argus II implantation, and had an average increase in visual cortical thickness of 2.2 percent relative to the pre-implantation scan (Figure S5B). Patient A10 showed the largest increases in left Lingual Gyrus (5.0 percent), the left Cuneus Cortex (3.5 percent), and the right Lingual Gyrus (4.3 percent).

#### Multiple Post Implantation Scans (One Case Study)

Patient A2 was scanned two additional times following the first scan at 43.5 months (3.63 years) post-implantation. The second scan was at 57 months (4.75 years) post-implantation with the device still in active use. The final scan was at 106 months (8.83 years) post-implantation, but 36 months (3 years) after ceasing to use the device. The cortical thicknesses in the left and right Pericalcarine Cortex, Cuneus Cortex, and Lingual Gyrus decreased over one year of device use (between 3.63 years and 4.75 years). In all these regions except the left Pericalcarine Cortex, the cortical thickness re-bounded following the years of disuse (Figure S6A). The cortical thickness in the Lateral Occipital Cortex (left and right hemisphere) increased over one year of Argus II use (similar to the increase in thickness in the group level analysis). The increased cortical thickness in the left and right Lateral Occipital Cortex was maintained after disuse of the Argus II device. The left and right LGN volumes are reported in the Supplemental Information (Figure S6B).

## Discussion

### Overview

Multiple early visual regions had significantly reduced cortical thickness or volume in the blind control group relative to the sighted control group, including the left Cuneus cortex, the left Lateral Occipital Cortex, the right Lingual Gyrus, and the right LGN. Similar to the blind control group, the Argus II patient group also had a significantly reduced cortical thickness in the right Lingual Gyrus and a significantly reduced volume in the left and right LGN relative to the sighted control group. However, the Argus II patient group had a significantly thicker left Cuneus Cortex and left Lateral Occipital Cortex relative to the blind patient cohort, and these thicknesses were similar in magnitude to the sighted control group. The Argus II patients cortical thickness across all left visual regions significantly partially correlated with the duration since implantation. Furthermore, the pre and post implantation case studies show no cortical thickening with a half year of use, but thickening with nearly four years of use. In another case study, the patient had an increase in thickness in the left and right Lateral Occipital Cortex between 3.63 and 4.75 years of device use, and a maintenance of that increased thickness following 3 years of disuse.

These results show that the Argus II patients can have a significant restoration of visual grey matter with artificial vision, especially following prolonged use and in visual cortical regions. As would be expected with structural changes in the brain, this rejuvenation of thickness increased with longer device usage, and appeared to require years of rehabilitated vision to be fully instantiated.

### Vision Loss and Cortical Thinning

Visual impairment due to retinal disease in the late blind has been shown to reduce the cortical thickness of visual regions (as discussed in the Introduction section). Cortical grey matter loss in patients with retinal disease is hypothesized to originate in the retina and gradually progress up the visual pathway via trans-synaptic neurodegeneration called Wallerian Degeneration (Machado and others 2017). At the cellular level, grey matter loss is hypothesized to be due to “neuronal apoptosis, variations in cortical myelination, alterations of the synaptic complexity, or a summation of these events” (Sanda *et al*. pg. 3474) (Burge and others 2016; Sanda and others 2018; Wagstyl and others 2015; Zilles and Amunts 2015).

In this paper we investigated late-blind Retinitis Pigmentosa patients, which have been shown to have reduced cortical thickness in visual regions. In particular, Castaldi *et al*. found significant thinning of V1 in RP patients compared to sighted controls (Castaldi and others 2019), however, this difference may be due to the younger age of the sighted controls. In Sanda *et al*. Retinitis Pigmentosa patients with tunnel vision (10-20 degrees) were found to have reduced cortical thickness in early visual cortex, V1 and V2, as well as, V3d, and V4 (Sanda and others 2018). Machado and colleagues studied the total grey matter volume in visual cortical regions, and found that the left and right Calcarine Sulci, left and right Lingual Gyri, left and right Cuneus, and right Superior Gyrus of the Occipital lobe were significantly reduced in volume in RP patients relative to sighted controls (Machado and others 2017). In contrast to these results, Ferriera et al found no significant differences between the cortical thickness of the RP patients they evaluated and their sighted controls (Ferreira and others 2017). Interestingly, when they only evaluated the region of primary visual cortex with spared vision, they found that RP patients with less than 15 degree visual field had a thicker visual cortex than the RP patients with more than a 15 degree visual field. This thickening of the more impaired patients may be due to compensation in regions of spared vision. This contrast of thinning in regions of impairment and thickening in regions of spared vision may contribute to the intersubject variability in RP patients.

The late-stage RP patients studied in this paper (*i.e.,* the blind control group) had severe vision loss, ranging from complete blindness (no light perception) to 2 degrees of tunnel vision remaining. Similar to results in other studies, we found significant degradation of grey matter in these RP patients from visual subcortical regions (such as LGN) to visual cortical regions (such as the Cuneus Cortex). An interesting difference between this study and Sanda et al. is that our RP patient cohort did not have a significantly thinner pericalcarine cortex (or early visual cortex) than the sighted controls. While the RP patients in our study had on average a substantially thinner pericalcarine cortex relative to the sighted control participants, this difference was not significant due to a large variability within RP patient group. This high variability in the cortical structure and function of low vision and blind individuals is common in the neuroscience literature (Cunningham and others 2015b; Ferreira and others 2017) and could in this case be caused in part by the conflicting processes of thinning and thickening of V1 during the progression of RP (as highlighted above and in (Ferreira and others 2017). Overall, the RP patient cohort in this study had significant grey matter loss (cortical thickness and subcortical volume) in multiple visual regions, which is expected due to the severity and long-duration of vision loss in the patients tested.

### Cortical Thickness Changes with Vision Restoration

Cortical thickness has been previously evaluated in two vision restoration populations: Argus II patients, and cataract patients. The two previous Argus II patient studies, which evaluated cortical thickness, were case studies (Cunningham and others 2015a; Stanga and others 2021). Cunningham *et al*. evaluated the V1 cortical thickness of two Argus II patients with RP (duration of prosthesis use was 1.5 months and 3.75 months) relative to late-stage RP controls (*N* = 9) and sighted controls (*N* = 9) (Cunningham and others 2015a). They found that the two Argus II patients’ V1 cortical thicknesses were within the ranges of the blind and sighted control groups, and argued that MRI imaging was feasible with the Argus II retinal prosthesis. The Cunningham patients were different than our patient group in two key aspects, Cunningham et al did not have pre-implantation and post-implantation within-subject comparisons, and their patients used the Argus II device for less than 4 months. Our analyses included two pre and post implantation case studies, one longitudinal post-implantation case study, and seven additional Argus II patients with a range of use from 19 months to 121 months.

Stanga *et al*. measured the cortical thickness of V1 and V2 in one Age-Related Macular Degeneration patient before and 13 months after implantation with the Argus II device (Stanga and others 2021). The cortical thickness of the Argus II patient’s V1 and V2 increased after implantation but was still below the age-matched sighted controls (*N* = 8) average cortical thickness. Stanga *et al*. studied an AMD patient with potentially substantial remaining visual perception, therefore their result is not highly comparable to our study in RP patients with light perception or less. However, they do show visual region rejuvenation with Argus II device use, which is generally consistent with our results.

Guerreiro *et al*. studied cortical thickness changes in patients with sight recovery due to cataract surgery (Guerreiro and others 2015) in patients with congenital cataracts. Cataract surgery was performed in these patients between 5 and 24 months from birth, and therefore within the visual critical period. Early blind patients with vision loss during the critical period have been previously found to have thicker early visual cortices, which is hypothesized to be due to an absent or reduced cortical pruning phase of visual development (Guerreiro and others 2015). Guerreiro found that early-blind patients with vision restoration following cataract extraction maintained this thickened visual cortex relative to sighted controls. Therefore, the decades of restored visual perception did not reverse the changes in cortical architecture due to blindness during the critical period. The critical period is a developmental phase with strong structural plasticity, and it is followed by adulthood with more limited plasticity. Our patient group were late blind and therefore did not have vision loss during the critical period but rather during adulthood. It is therefore possible that while the structural changes due to impairment during the critical period cannot be reversed in congenital cataract patients, our results show a potential to partially reverse changes that can occur in adulthood in Argus II patients.

Our group results show that two later visual regions (the left Cuneus Cortex and Lateral Occipital Cortex) had significant rejuvenation in Argus II patients that was equivalent to the sighted controls. Furthermore, correlation analyses between visual cortical region thickness and Argus II patients duration of use, as well as case studies fortify our group result by showing more rejuvenation with longer device use. If it is the case that visual cortical thickness is increased with vision restoration, we would hypothesize the process is a reversal of the previously studied Wallerian degeneration (discussed above). In contrast to the degeneration process, during the rejuvenation process we hypothesize that the surviving visual neurons increase their myelination and synaptic and dendritic complexity.

A remaining question is the relationship between this cortical rejuvenation and the behavioral outcomes with artificial vision. In other words, does the cortical rejuvenation impact the level of functionality of patients with the Argus II device? Our behavioral measure of visual shape matching did not significantly correlate with visual cortical thickness in the Argus II patients. However, this nuanced function-structure relationship likely either requires a larger variety of behavioral tasks (as Argus II patients functionality can vary depending on task) or a larger patient cohort to be fully elucidated.

We next discuss the two regions which had significant rejuvenation in our group level analysis, the Lateral Occipital Cortex and the Cuneus Cortex, and their potential relationship to visual function in the visually restored.

### The Lateral Occipital Cortex and Vision Restoration

The Argus II patients’ thicker left Lateral Occipital Cortex relative to late blind controls (and similar to sighted controls) is likely due to the restored visual function from the artificial vision. While the comparison between participant groups is not causal (*i.e.,* a pre and post measure), it does indicate a significant difference relative to blind controls with the same retinal disease (Retinitis Pigmentosa), comparable ages, and a similar level of natural vision loss.

Lateral Occipital Cortex (Figure 2A) in the Desikan-Killiany atlas covers multiple sulci and gyri within the occipital lobe (description in the Klein and Tourville Appendix (Klein and Tourville 2012)) including portions of the Transverse Occipital Sulcus, and the Lateral Occipital Sulcus. The Lateral Occipital Cortex is the highest level in the visual processing hierarchy within the occipital lobe in the Desikan-Killiany atlas, in particular, the visual regions progress from the Pericalcarine Cortex (including most of the Primary Visual Cortex) up to the Lateral Occipital Cortex (including mostly V2 and V3). The Lateral Occipital Cortex has been shown (using neuroimaging techniques) to process objects’ three dimensional shape (Doniger and others 2000; Grill-Spector and others 2001). Furthermore, patients with lesions in the Lateral Occipital Cortex have been found to have deficits in the recognition of object shape (Moscovitch and others 1997; Ptak and others 2014). In addition, Transcranial Magnetic Stimulation (TMS) that suppresses the activity of the Lateral Occipital Cortex can generate a deficit in the object recognition process (Stewart and others 2001). Recent TMS research points to a feedback of perceived object size information (in contrast to earlier measures of retinal size) from the Lateral Occipital Cortex to early visual cortex (Zeng and others 2020). The Lateral Occipital Cortex has also been shown to process multisensory information, in particular, integrating shape information between tactile, visual, and auditory modalities (Amedi and others 2007; Beauchamp 2005).

Argus II patients have been shown to be able to recognize basic objects (Luo and others 2014) and integrate shape across the senses (Stiles and others 2022) with artificial vision. Therefore, the reinvigoration of the Lateral Occipital Cortex could be an important part of rehabilitating these basic visual functions with artificial vision. Furthermore, the Lateral Occipital Cortex’s direct feedback connections to earlier visual cortices (shown in previous TMS experiments (Zeng and others 2020)) may have a critical role in the early processing of visual and multisensory information in the visually restored brain. In general, the hypothesis that a thickened left Lateral Occipital Cortex could facilitate the rehabilitation of Argus II patients is consistent with the results in this paper and in the literature.

### The Cuneus Cortex and Vision Restoration

The Argus II patients also had a significantly thicker left Cuneus Cortex relative to blind controls. The Cuneus Cortex (Figure 2A) in the Desikan-Killiany atlas covers multiple sulci and gyri within the occipital lobe (description in the Klein and Tourville Appendix (Klein and Tourville 2012)) including portions of the Parieto-occipital Sulcus, and the posterior limit and dorsomedial margin of the Calcarine Sulcus. The Parieto-occipital Sulcus within the Cuneus Cortex has been shown (using neuroimaging techniques) to process mental navigation and egocentric spatial tasks in humans and non-human primates (Ino and others 2002; Pitzalis and others 2021). A lesion study in non-human primates connects the Parieto-occipital Sulcus with visuomotor control, in particular lesions generated deficits in reach and grasp tasks (Battaglini and others 2002; Galletti and others 2003). Therefore, whereas the Lateral Occipital Cortex is in the ventral stream and processes the identity of objects, the Cuneus Cortex is located in the dorsal stream and is attuned to the location and orientation of the body relative to objects. The Argus II patients are trained to navigate with their devices and have shown the utility of visual information in mobility tasks (Flourence and others 2023; Jeganathan and others 2022; Sadeghi and others 2024). Therefore, the thickening of the Cuneus Cortex observed in this study is consistent with the functionality measured in Argus II patients.

### Conclusions

Overall, the Argus II patients were shown to have a thicker cortex relative to the blind controls in two higher visual regions (the Cuneus Cortex and the Lateral Occipital Cortex). A correlation analysis showed that longer using Argus II patients had on average a thicker visual cortex and two pre and post implantation case studies fortified this trend. Therefore, these results support the theory that the visual cortex can be structurally rejuvenated with vision restoration following decades of late blindness and visual cortical atrophy.

## Data Availability

Neuroimaging data will be made available through the Connectome Coordination Facility (CCF) (https://www.humanconnectome.org/).

## Acknowledgments

We are grateful for support from the National Institutes of Health, National Eye Institute (1U01EY025864-01); the National Institutes of Health, the BRAIN Initiative (5K99EY031987-02); the Arnold O. Beckman Postdoctoral Scholars Fellowship Program; the USC Roski Eye Institute; and the Philanthropic Educational Organization Scholar Award Program.

