## Supplemental Information for "Visual Cortical Thickness Increases with Prolonged Artificial Vision Restoration"

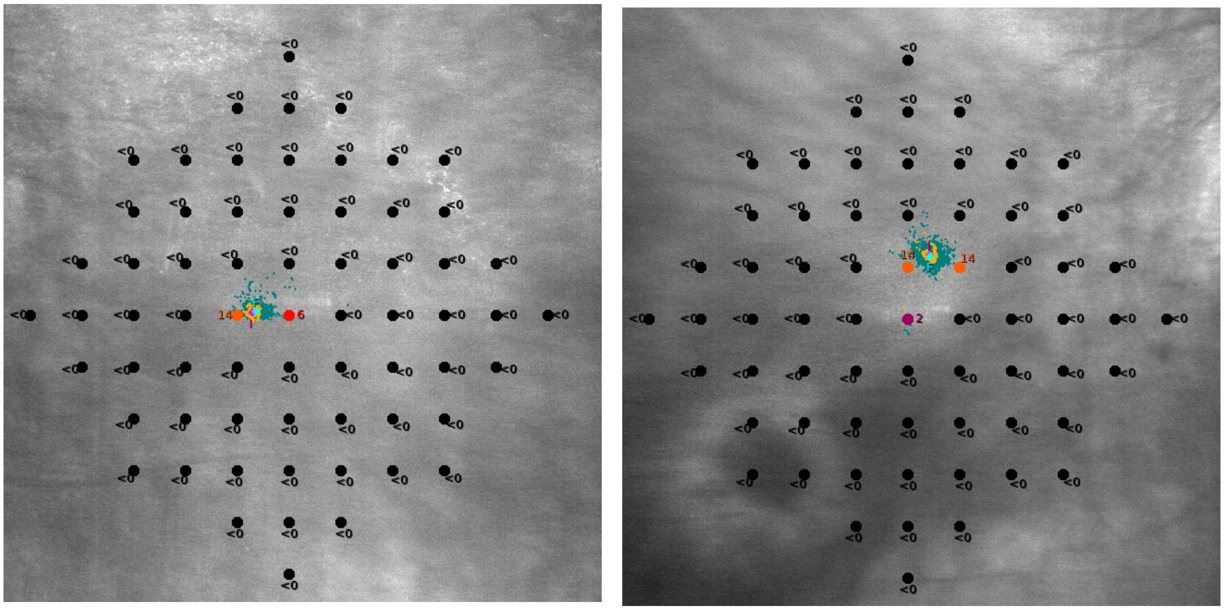

**Left Eye (OS)** **Right Eye (OD)**

**Figure S1: MAIA Microperimetry Results for Patient B6.** Patient B6 had visual acuity of 20/80-2 in the left eye and 20/50+2 in the right eye (Table 1). However, this patient was classified as severe RP by the severe limitation to their visual field (2 degrees in both eyes). The microperimetry data in Figure S1 shows this visual field, which was tested with a custom grid pattern, 4-2 strategy, and 200 millisecond stimuli duration on a MAIA machine. The stimuli were presented 2 degrees apart (laterally and vertically) and the colors and numbers in the images represent the threshold frequency (dB); green represents good visual light perception (normal range), and black represents no light perception detected. Patient B6 shows intermediate to low visual light detection with a range of 14 dB to 2 dB (the orange to red dots in the images) in two to three locations in each eye, with the rest of the visual field containing no light detection.

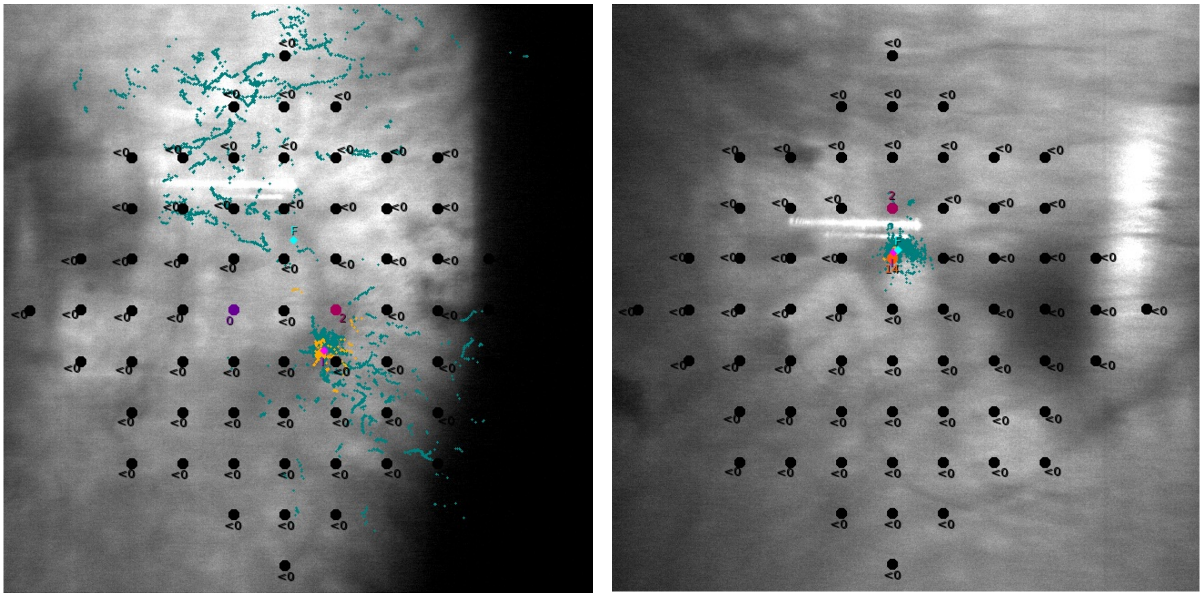

**Left Eye (OS)** **Right Eye (OD)**

**Figure S2: MAIA Microperimetry Results for Patient B9.** Patient B9 had visual acuity of 20/800 in the left eye and 20/300+1 in the right eye (Table 1). This patient was classified as severe RP by both the low visual acuity and by the severe limitation to their visual field (2 degrees in both eyes). The microperimetry data in Figure S2 shows this visual field, which was tested with a custom grid pattern, 4-2 strategy, and 200 millisecond stimuli duration on a MAIA machine. The stimuli were presented 2 degrees apart (laterally and vertically) and the colors and numbers in the images represent the threshold frequency (dB); green represents good light perception (normal range), and black represents no light perception detected. Patient B9 shows intermediate to low visual light detection with a range of 14 dB to 2 dB (the orange to red dots in the images) in two to three locations in each eye, with the rest of the visual field containing no light detection (0 dB or less than 0 dB). The MAIA machine classified Patient B9 as having unstable fixation in the left eye (scattered teal lines in left image shows fixation irregularity), and stable fixation in the right eye (the teal lines and dots are clustered at fixation).

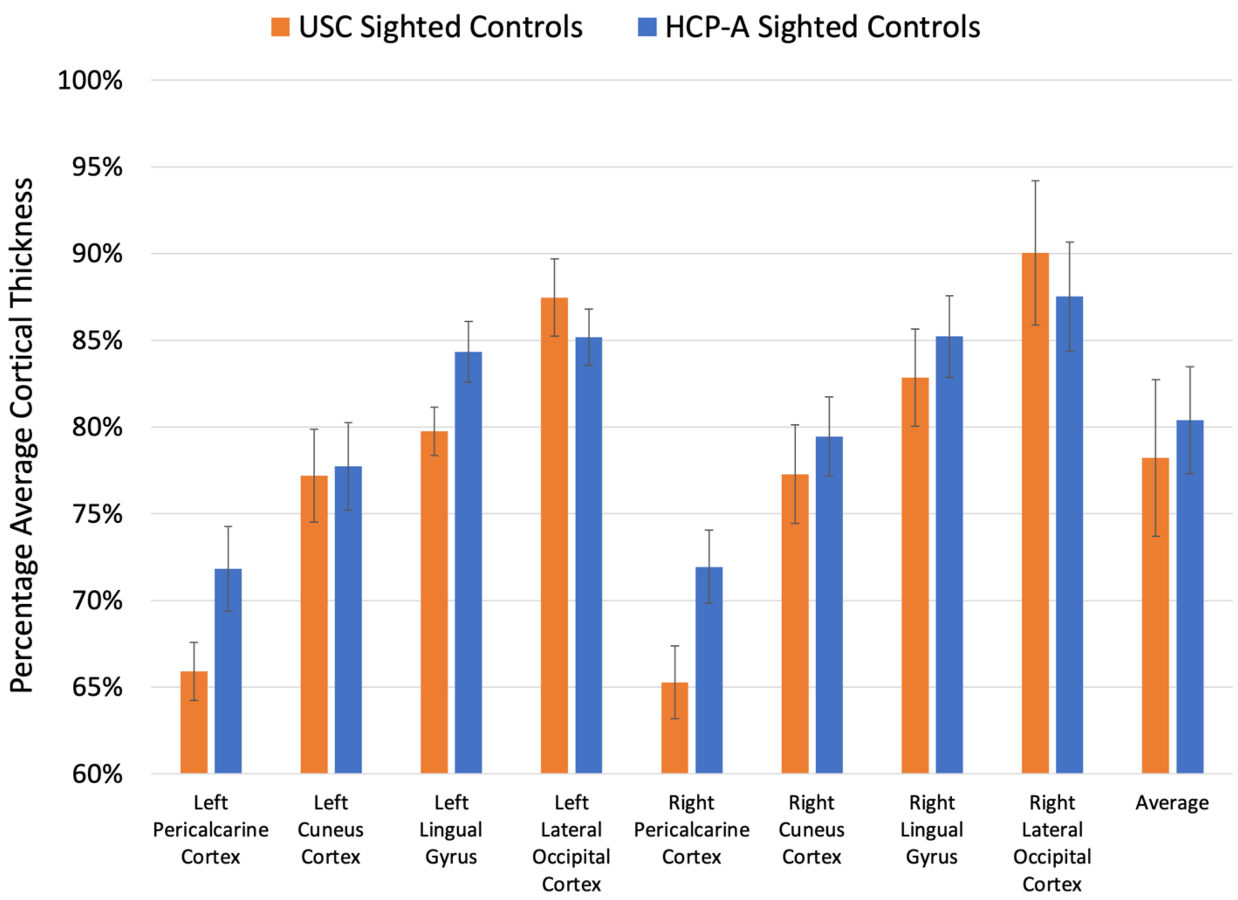


**Figure S3: Comparison Between USC and HCP-A Sighted Controls.** The sighted controls MRI data were collected at either USC or as a part of the HCP-A protocol. When the data from these two different sites were compared with a Wilcoxon rank sum test (one for each cortical region on the left and right) there was not a significant variation between the USC sighted and the HCP-A sighted controls ($p > 0.05$) except for the left pericalcarine cortex ($p = 0.04$) and left lingual gyrus ($p = 0.04$). The full length of each error bar represents one standard deviation.

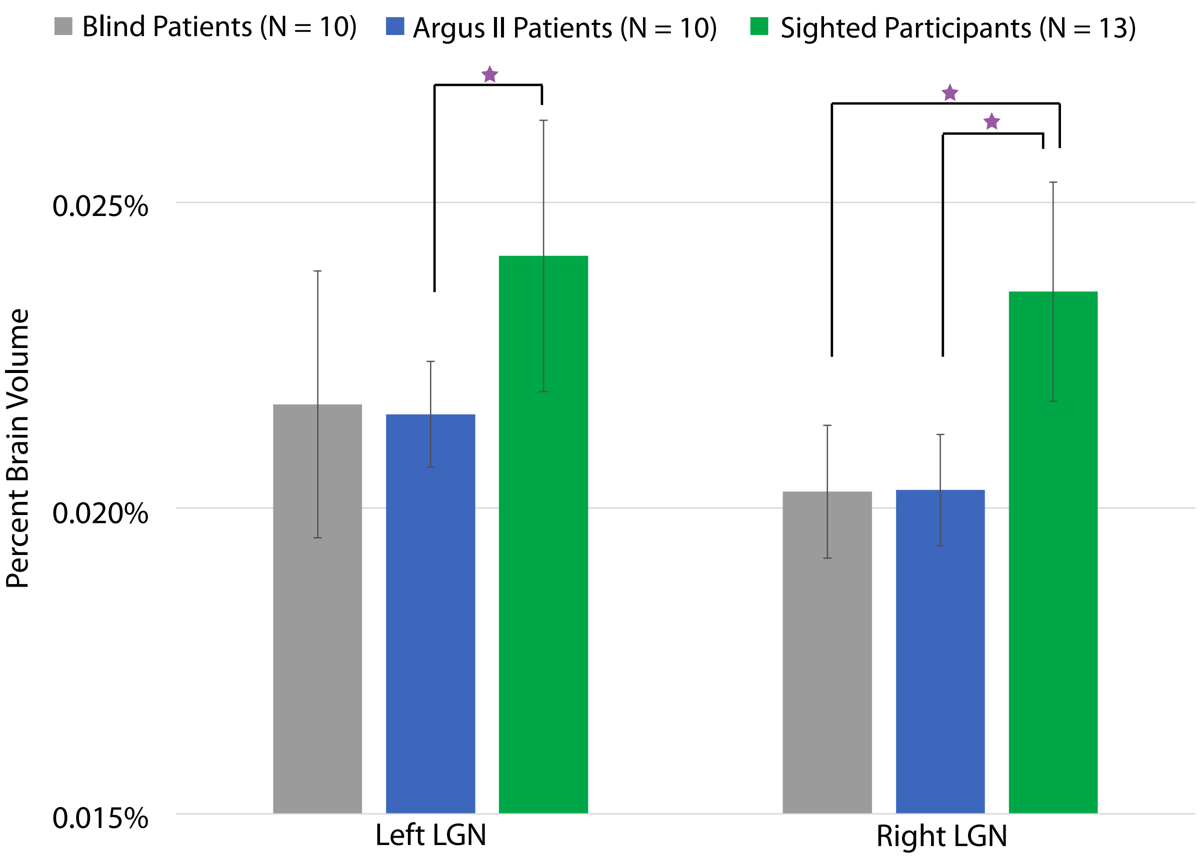


**Figure S4: Lateral Geniculate Nucleus Volume.** Figure S4 shows the Lateral Geniculate Nucleus (LGN) volume for the three participant groups: the blind patients, the Argus II patients, and the sighted controls (details in Table 1). The left and right LGN volumes are normalized by the participant's brain segmentation volume without ventricles. Purple stars indicate significant differences between participant groups ($p < 0.05$). The full length of each error bar represents one standard deviation. The left LGN volume had significant variation across the participant groups ($H\,(2,\ n = 33) = 6.7$, $p = 0.04$). The group-wise comparisons showed that the sighted control group had a significantly larger right LGN than the Argus II patient group (Argus II patients vs. Blind controls: $p = 0.62$; Argus II patients vs. Sighted controls: $p = 0.02$; Sighted controls vs. Blind controls: $p = 0.06$). The right LGN volume had significant variation across the participant groups ($H\,(2,\ n = 33) = 9.16$, $p = 0.01$). The group-wise comparisons showed that the sighted control group had a significantly larger right LGN than the Argus II patient group and the blind patient group (Argus II patients vs. Blind controls: $p = 0.92$; Argus II patients vs. Sighted controls: $p = 8.8 \times 10^{-3}$; Sighted controls vs. Blind controls: $p = 0.01$).

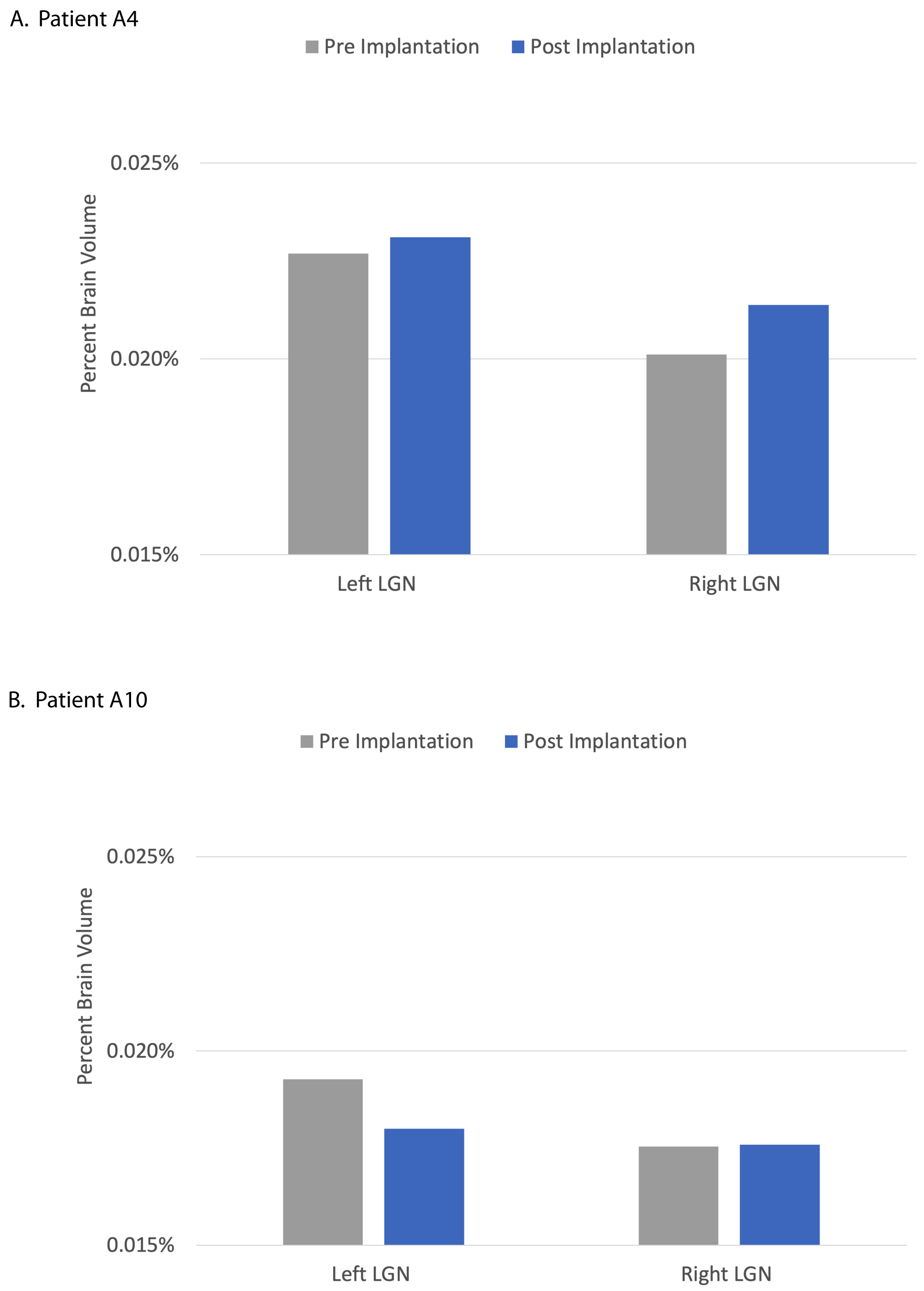


**Figure S5: Individual Pre and Post Implantation LGN Volumes.** Figure S5 shows the volumes of the left and right LGN for two case studies (Patients A4 and A10), which have scans before and after the implantation of the Argus II device (details in Table 1 and the Methods and Results section). Patient A4 was scanned 6.5 (0.54 years) after Argus II implantation and Patient A10 was scanned 44.5 months (3.71 years) after Argus II implantation

A.

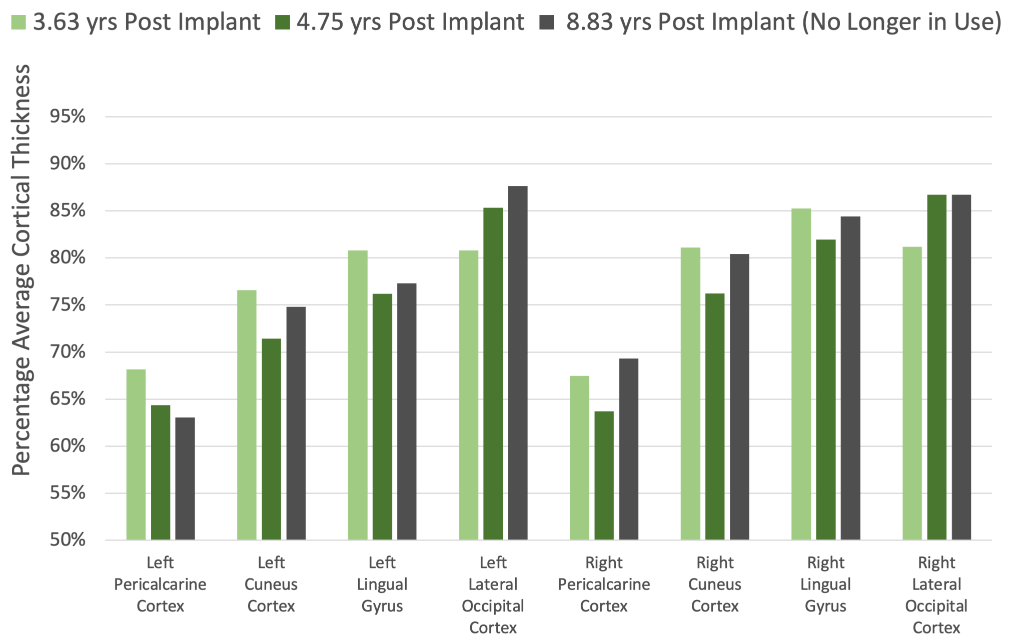


B.

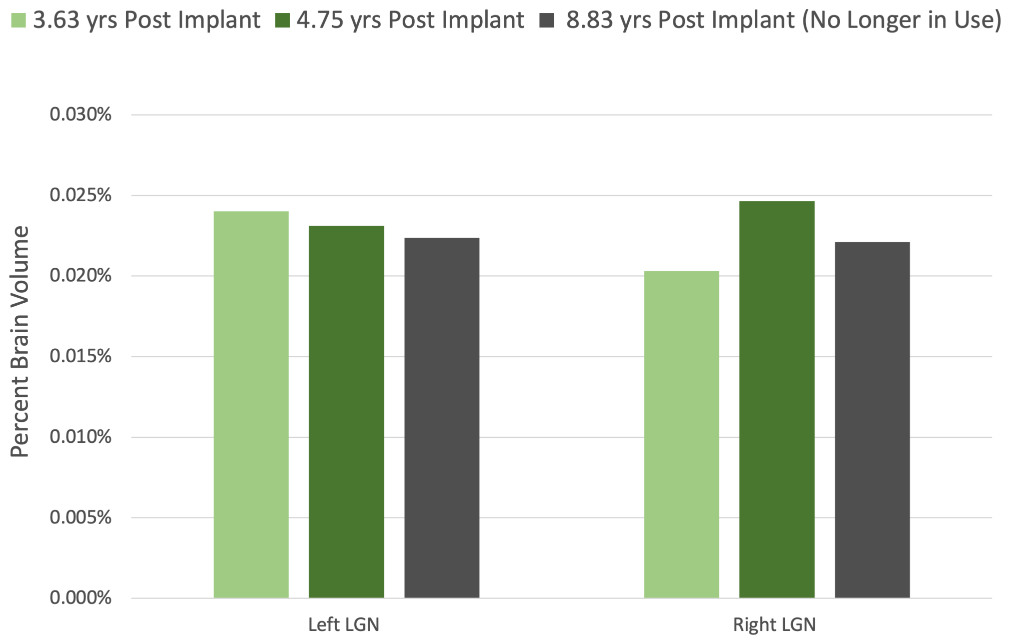


**Figure S6: Post Implantation Longitudinal Data.** Figure S6 shows the visual cortical thicknesses and the volumes of LGN for one longitudinal case study (Patient A2), which has scans at three time points after the implantation of the Argus II device (Details in the Methods section and Results section).

| Subject ID | Participated in rehabilitation training? | Total rehabilitation training hours | Did the patient visit a University or Second Sight for additional testing? * | Frequency of device use (averaged over total time since implant) |
|---|---|---|---|---|
| **A1** | Yes | 2 hours | Yes | Once per week |
| **A2** | Yes | 12 hours | Yes | Once per day |
| **A3** | Yes | 24 hours | Yes | Every other day |
| **A4** | - | - | Yes | - |
| **A5** | Yes | 30 hours | Yes | Once per week |
| **A6** | Yes | 13 hours | Yes | Every day |
| **A7** | Yes | 150 hours | No | Once per day |
| **A8** | No | - | Yes | Once per week |
| **A9** | No | - | Yes | Once per month |
| **A10** | Yes | 14 hours | No | Once per month |

** This excludes the University visit(s) for this research study;  - = Data not available or not applicable*

**Table S1. Argus II Training Information.** Self-reported Argus II patient directed rehabilitation training and use. Column two reports whether patients participated in the Second Sight provided rehabilitation training. Second Sight training duration (estimated by the patient) is reported in column three. The visits to universities for research studies that can have similar results to directed training are reported in column 4. Patients reported that they used the Argus II device in their home between every day and once per month as reported in column 5π. Patients' duration of prosthesis use is reported in Table 1.
